# Automated Prognosis of Renal Function Decline in ADPKD Patients using Deep Learning

**DOI:** 10.1101/2023.01.13.23284471

**Authors:** Anish Raj, Fabian Tollens, Anna Caroli, Dominik Nörenberg, Frank G. Zöllner

**Affiliations:** Computer Assisted Clinical Medicine, Medical Faculty Mannheim, Heidelberg University, Mannheim, 68167, Baden Württemberg, Germany; Mannheim Institute for Intelligent Systems in Medicine, Medical Faculty Mannheim, Heidelberg University, Mannheim, 68167, Baden Württemberg, Germany; Department of Radiology and Nuclear Medicine, University Medical Centre Mannheim, Medical Faculty Mannheim, Heidelberg University, Mannheim, 68167, Baden Württemberg, Germany; Bioengineering Department, Istituto di Ricerche Farmacologiche Mario Negri IRCCS, Ranica (BG), 24020, Italy

**Keywords:** Autosomal dominant polycystic kidney disease, deep learning, total kidney volume, chronic kidney disease, classification, prediction

## Abstract

The prognosis of renal function decline in Autosomal Dominant Polycystic Kidney Disease (ADPKD) is vital for early intervention. Currently, the accepted biomarkers are height-adjusted total kidney volume (HtTKV) with estimated glomerular filtration rate (eGFR) and patient age. However, kidney volume delineation is time-consuming and prone to observer variability. Furthermore, improvement in prognosis can be achieved by incorporating automatically generated features of kidney MRI images in addition to the conventional biomarkers. Hence, to improve prediction we develop two deep learning algorithms. At first, we create an automated kidney volume segmentation model that can accurately calculate HtTKV. Secondly, we use the segmented kidney volumes with the predicted HtTKV, age, and eGFR at the baseline visit. Here, we use a combination of convolutional neural network (CNN) and multi-layer perceptron (MLP) for the prediction of chronic kidney disease (CKD) stages >=3A, >=3B, and a 30% decline in eGFR after 8 years from the baseline visit. We obtain AUC scores of 0.96, 0.96, and 0.95 for CKD stages >=3A, >=3B, and 30% decline in eGFR, respectively. Moreover, our algorithm achieves a Pearson correlation coefficient of 0.81 between predicted and measured eGFR decline. We further extend our approach to predict distinct CKD stages after eight years with high accuracy. The proposed approach might improve monitoring and support the prognosis of ADPKD patients from the earliest disease stages.

## 1. Introduction

Autosomal dominant polycystic kidney disease (ADPKD), due to the growth of cysts and therefore, degeneration of renal parenchyma leads to end-stage renal disease (ESRD) and renal failure [1]. It affects up to 12 million people worldwide. ADPKD accounts for up to 10% of patients with ESRD [1]. Given these numbers, early treatment of the disease when the renal parenchyma is still preserved is warranted. However, disease progression monitoring is difficult since kidney function may remain normal for several decades and is therefore not informative in the earliest stages of the disease. Clinical, genetic, environmental, epigenetic, and radiologic factors have been studied as predictors of progression to kidney failure in ADPKD [2, 3]. On the other hand, kidney volume has been shown to be a promising marker for disease progression [4, 5]. It is also recognized by the Food and Drug Administration (FDA) as a candidate for a prognostic biomarker for ADPKD progression [6] and used within recent clinical phase 3 studies (TEMPO 3/4 trial, primary outcome measure; the annual rate of change in TKV over time) [7, 8].

Magnetic resonance imaging (MRI) is recognized as an important tool to monitor the progression of ADPKD, and the total kidney volume (TKV) or the height-adjusted total kidney volume (HtTKV) has been shown to predict renal function decline in ADPKD patients [9, 2].

Thereby, a complex interaction of prognostic factors like clinical, genetic, environmental, epigenetic, and radiologic determines the number of kidney cysts and their growth rates, which affect total kidney volume (TKV)[2]. Based on this, prognostic models like the Mayo imaging classification tool [9] have been developed to stratify ADPKD patients into classes and predict disease progression. A multiple linear regression model is generally employed for this task. However, Kline et al. [10] have shown that using the conventional biomarkers (HtTKV, age, and eGFR) may not be sufficient for accurate predictions. They reported that for predicting eGFR decline after 8 years, a Pearson correlation coefficient ‘r’ of only 0.51 could be achieved. They improved this prediction model by further incorporating texture features (repeating patterns of local image intensity variations that provide information regarding the spatial arrangement of image intensities) from T2-weighted MRI images. The texture features are extracted from manually segmented kidneys and consist of energy, entropy, and correlation features. By adding texture features to conventional biomarkers, the resulting Pearson’s r reaches −0.70. However, this approach, requiring manual segmentation of the kidneys, is time-consuming and observer-dependent. Moreover, automated kidney segmentation approaches are steadily proposed and are emerging into different applications in renal MRI [11, 12]. Considering this, we created a fully automated system that can segment the kidneys automatically and make an accurate prognosis. More in detail, we first develop a deep learning pipeline that segments kidneys from T2-weighted MRI and calculates HtTKV from the kidney segmentations. We then extract features from the segmented kidneys and combine them with the conventional biomarkers to predict renal function decline after 8 years, using data from 135 ADPKD patients with normal kidney function at baseline. Specifically, we target the following prognosis tasks: a) whether a patient will reach CKD stage 3A or not (eGFR < 60 ml/min/1.73 m^2^), reach 3B or not (eGFR < 45 ml/min/1.73 m^2^), and reach a 30% decline in eGFR or not, b) we classify each patient into a distinct CKD stage (CKD stages 1, 2, 3A, 3B, and 4) and c) predict percent change in the eGFR.

## 2. Methods

### 2.1. Patient Data

The patient data were acquired from the National Institute of Diabetes and Digestive and Kidney Disease (NIDDK), National Institutes of Health, USA and were recorded in the Consortium for Radiologic Imaging Studies of Polycystic Kidney Disease (CRISP) study[4]. The dataset consists of 241 patients in total. However, for our work, we selected patients who underwent T2-weighted MR imaging and had eGFR values > 70 ml/min/1.73 m^2^ at the baseline visit. These criteria were selected in accordance with [10, 9]. The resulting dataset contains information from 135 patients. Table 1 list the number of patients’ CKD stages at baseline and after 8 years alongside demographic data, eGFR, and kidney volumes.

**Table 1:** Demographic, clinical and radiological data of the 135 ADPKD patients included in the study. Data are shown as mean ± SD or Number (%)

|  | Baseline | 8-year Follow-up |
| --- | --- | --- |
| TKV (ml) | 974±585 | 1531±1135 |
| HtTKV (ml/m) | 559±322 | 880±615 |
| eGFR (ml/min/1.73 m<sup>2</sup>) | 96±25 | 76±28 |
| CKD stage 1 | 66 | 46 |
| CKD stage 2 | 69 | 44 |
| CKD stage 3A | - | 24 |
| CKD stage 3B | - | 14 |
| CKD stage 4 | - | 7 |
| > 30% eGFR decline | - | 48 |
| Gender (M/F) | 58/77 | - |
| Age (years) | 32±9 | - |

To allow for a comparison to the SOTA method [10], the patients were regrouped into two groups based on the presence or absence of each of the following conditions at 8 years after baseline: reaching CKD stage 3A (eGFR < 60 ml/min/1.73 m^2^), reaching CKD stage 3B (eGFR < 45 ml/min/1.73 m^2^), and 30% decline in eGFR. As depicted in table 2, our data resembles a similar class distribution as in the work of Kline et al. [10].

**Table 2:** Ratio of the positive number of samples to the total number of samples in our dataset as compared to the one in state-of-the-art (SOTA) approach from Kline et al. [10]. In all three criteria, we have more imbalance as compared to the work from Kline et al. [10].

| Criteria | current study | SOTA method |
| --- | --- | --- |
| Reached CKD stage 3A | 0.333 | 0.360 |
| Reached CKD stage 3B | 0.155 | 0.180 |
| 30% eGFR decline | 0.355 | 0.385 |

### 2.2. Data Pre-processing

We first normalized the renal MRIs, predicted HtTKV (ml/m) (obtained as per section 2.3.1), age (years), and eGFR (ml/min/1.73 m^2^) at the baseline visit using the Z-score normalization equation (1),

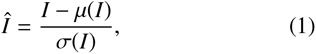

where *I* and *Î* are the original and normalized image/feature, respectively. The T2-weighted volumes were recorded with a size of 256 × 256 pixels and 12-30 slices [4]. However, the network for prognosis requires uniformly sized image volumes as inputs. Hence, we center-cropped and/or padded (as required) the image volumes to the size of 224 × 224 × 16 voxel. We ensure that complete kidney volumes were present within this reshaped volume.

### 2.3. Deep learning models

We implement two deep learning models for a fully automatic prognosis. We first used a segmentation network called attention U-Net (figure 1) [15] to extract kidneys from MRI volumes. Thereafter, we use the segmented kidney volumes as input to our prognosis network (figure 2) to make the final prognosis. We implement this two-step process so that we can first derive HtTKV automatically and then use the output from the first network and feed it to the second network for the classification/regression task.

**Figure 1:**
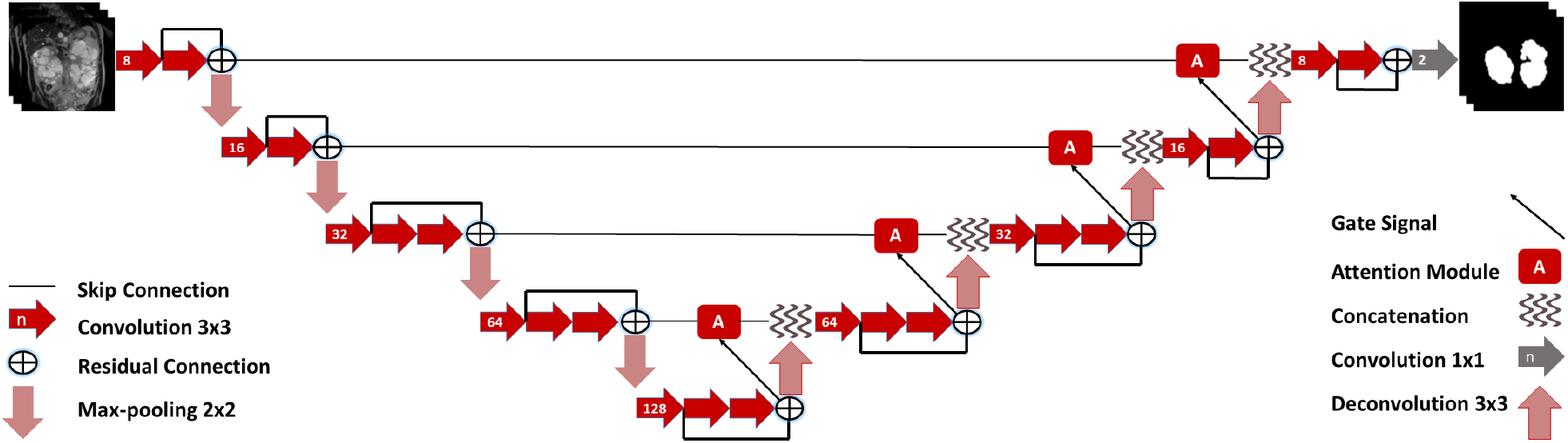
Segmentation network: Attention U-Net with attention modules. The network is U-Net based [13] with attention gates that help the network focus on relevant image regions, e.g., kidneys. The network is used to segment kidneys from patient MRI volumes (baseline visit). The segmented kidneys are then used to calculate the HtTKV of the patient. Adapted from [14, 15].

**Figure 2:**
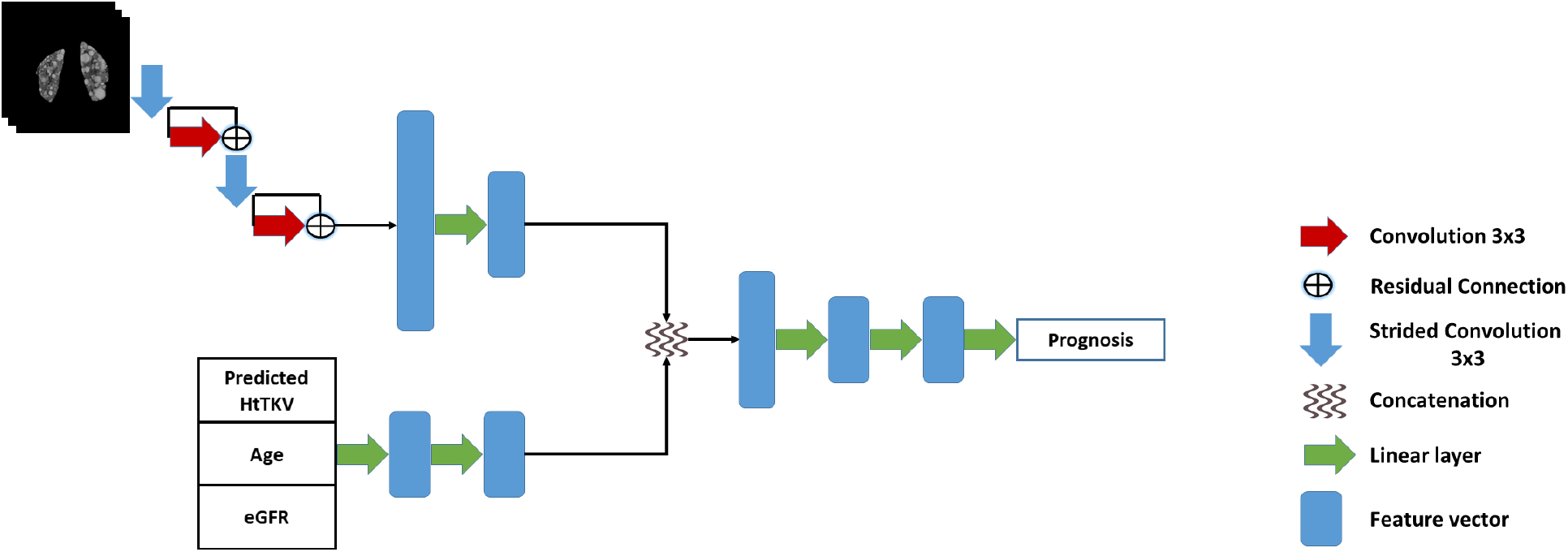
Our proposed prognosis network architecture for classification/regression prognosis. The first input is patient MRI volume (baseline visit) that contains only segmented kidneys obtained from the segmentation network. The convolution layers are used to extract features from the volumes. The second input consists of a vector of predicted HtTKV, age, and eGFR at the baseline visit. Here, an MLP extracts features. Then, the features from both inputs are concatenated and used to make the final prognosis. Every convolution, strided convolution, and linear layer is followed by instance normalization and PReLU activation [16]. The strided convolutions have a stride of 4. There is no final activation for the regression task, but for classification, we use sigmoid.

#### 2.3.1. Segmentation network

The first network is based on the attention U-Net [15]. We used this network to segment the kidneys from MRI volumes. Briefly, this network is composed of a Ushaped encoder-decoder architecture [13] and modified with attention gates [15], which help focus the network on relevant image regions. This is achieved by using higher-level features as a guide to suppress noise and irrelevant features in the lower-level features. We use the approach described in [14] to train and apply the attention U-Net. Initially, this network was trained and evaluated on T1-weighted MRI for renal segmentation. We extend this work by training the attention U-Net on T2-weighted images. The network architecture is illustrated in figure 1.

The network was trained on 100 T2-weighted volumes of our dataset for which manually segmented kidneys were available. Kidney annotations were done similarly as described in [14]. To obtain kidney segmentations for the remaining patients without manual annotation, the trained network was applied. All automatically segmented kidneys from the network are then used to calculate the HtTKV. The HtTKV can be calculated by multiplying the number of segmented kidney voxels by the voxel volume (in mm^3^) and then dividing it by 1000 and the height of the patient (in m) to get the volume in ml/m.

#### 2.3.2. Prognosis network

Our proposed network for predicting the final prognosis consists of two parts: a CNN and an MLP. The CNN part consists of convolutions and strided convolutions. The strided convolutions have a stride of 4, hence, each time, the spatial dimensions of the image/features are downsampled 4 times. The MLP part contains linear layers that produce one-dimensional feature vectors. The CNN part takes image volumes as input, while the MLP part accepts a vector of predicted HtTKV, age, and eGFR from the baseline visit. The features from CNN and MLP are concatenated and then a final MLP is used to make a prognosis. The network architecture is shown in figure 2. Each of the convolutional, strided convolutional, and linear layers are followed by an instance normalization layer [17] and PReLU activation [16] (except for the last linear layer). Depending upon the task type, the last layer is followed by the sigmoid (binary classification) or softmax (multi-class classification) activation in the case of classification tasks or no activation in the case of the regression task.

The prognosis after 8 years is divided into two tasks:

1. Classification: this is further divided into binary and multi-class classification tasks.
  a. Binary classifications: whether a patient will reach CKD stage 3A or not (eGFR < 60 ml/min/1.73 m^2^), reach 3B or not (eGFR < 45 ml/min/1.73 m^2^), and reach a 30% decline in eGFR or not.
  b. Multi-class classification: here, we classify each patient under a distinct CKD stage category after 8 years (CKD stages 1, 2, 3A, 3B, and 4).
2. Regression: percent change in the eGFR.

As illustrated in figure 2, we feed the segmented kidneys to a small convolutional neural network. Simultaneously, we use the predicted HtTKV, age, and eGFR at the baseline visit as input to a shallow multi-layer perceptron. Finally, we combine the features from kidney volumes and the MLP inputs and run them through a final MLP to make our prognosis.

### 2.4. Network implementations & training

#### 2.4.1. Segmentation network

The segmentation network for kidney segmentation is trained with a batch size of 16 and a patch size of 128. We used the adam optimizer [18] with a learning rate of 0.001. The loss function is cosine loss [14]. As the activation function, we employ exponential linear units (elus) [19] with batch-normalization, dropout (probability=0.01) and L2 normalization (10^−7^). Furthermore, we perform a 5-fold cross-validation and train for a minimum of 20 epochs. The train:validation:test split was 70:10:20 subjects in each fold. We employ early stopping to avoid overfitting and select the network with the highest dice score (validation data) for testing.

#### 2.4.2. Prognosis network

The prognosis network was trained for 30 epochs with 5-fold stratified cross-validation. The train:validation:test split was 95:13:27 subjects in each fold. The batch size and learning rate were 8 and 0.001, respectively. For classification tasks, we used weighted cross-entropy (CE) loss function and area under (AUC) the receiver operating characteristic curve (ROC) to select the best network. However, for the task of classifying distinct CKD stages, we use the f1-score to select the best network. We further use a weighted random sampling strategy to deal with the class imbalance in the classification of distinct CKD stages. Here, classes with less number of samples are weighted higher and accordingly sampled more often with replacement during training. Meanwhile, for regression, we implement mean squared error (MSE) loss and use the mean absolute error (MAE) score to choose the best network[20].

### 2.5. Evaluation

To evaluate and compare the results, we used the weighted f1-score (since we have a high class imbalance for CKD stage 4) and the AUC of the ROC for classification outputs. The weighted f1-score is defined as,

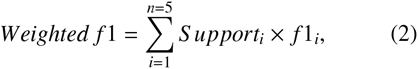

Where Support_*i*_ is the support proportion of each class *i*, given by,

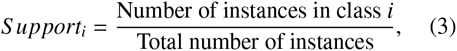

and precision, recall, and f1-score are given by

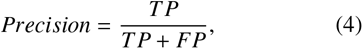

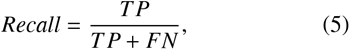

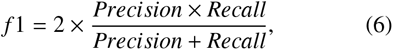

The TP, FP, and FN are true positives, false positives, and false negatives, respectively. In addition, we used Pearson’s correlation coefficient and Bland-Altman plots for evaluating regression results. Furthermore, for comparison, we employed the Mayo imaging classification tool [9, 21] to predict eGFR values at 8 years after baseline, using the baseline HtTKV.

## 3. Results

The results for T2-weighted kidney volume segmentations are displayed in figures 3 and 4. Furthermore, the results for the prognosis tasks are shown in table 3 and figures 5 and 6. The scatter plots (figures 3 and 6) also show Pearson’s correlation coefficient ‘r’ for better comprehension.

**Table 3:** Classification results of reaching CKD stage 3A, reaching CKD stage 3B and having a 30% eGFR decline after 8 years. Abbreviations: AUC = area under the curve, SOTA = state-of-the-art

| Criteria | Precision | Recall | AUC | SOTA AUC |
| --- | --- | --- | --- | --- |
| Reached CKD stage 3A | 0.951 | 0.866 | **0.965** | 0.940 |
| Reached CKD stage 3B | 0.900 | 0.857 | 0.957 | **0.960** |
| 30% eGFR decline | 0.846 | 0.916 | **0.952** | 0.850 |

**Figure 3:**
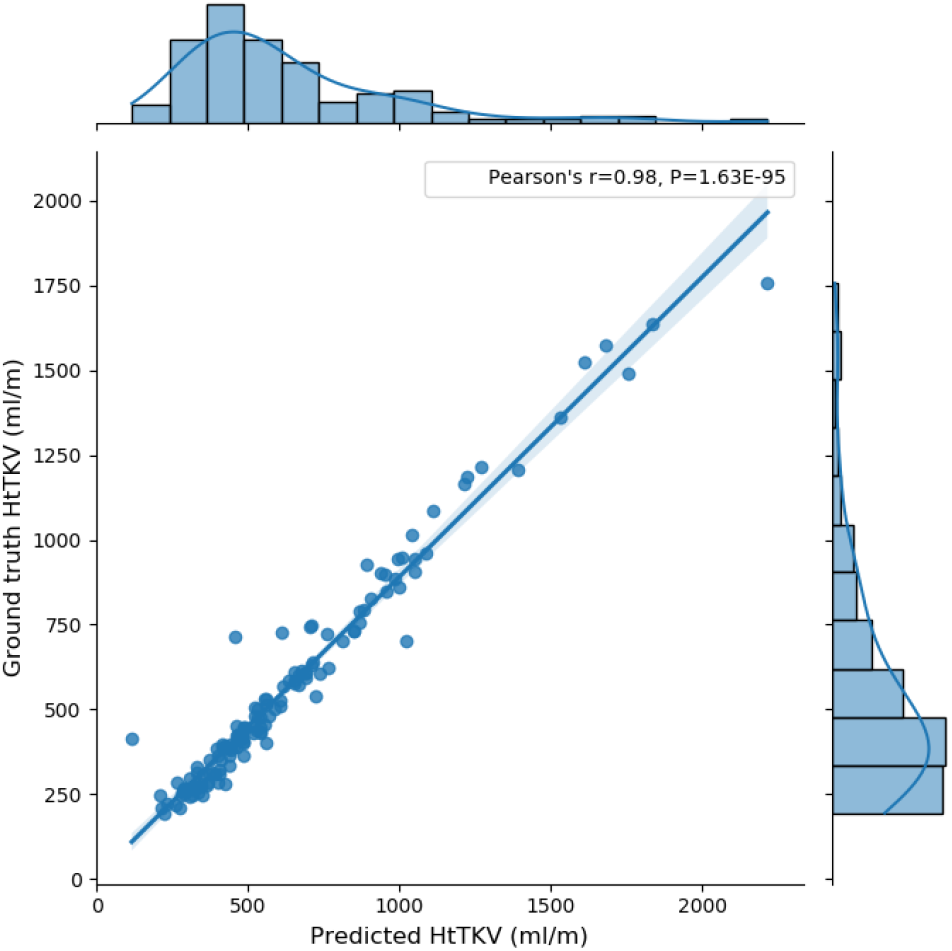
Baseline predicted HtTKV against the ground truth HtTKV. The predicted HtTKV is obtained from the segmentation network by segmenting kidneys from T2-weighted MRI volumes. The Pearson correlation coefficient is 0.98.

**Figure 4:**
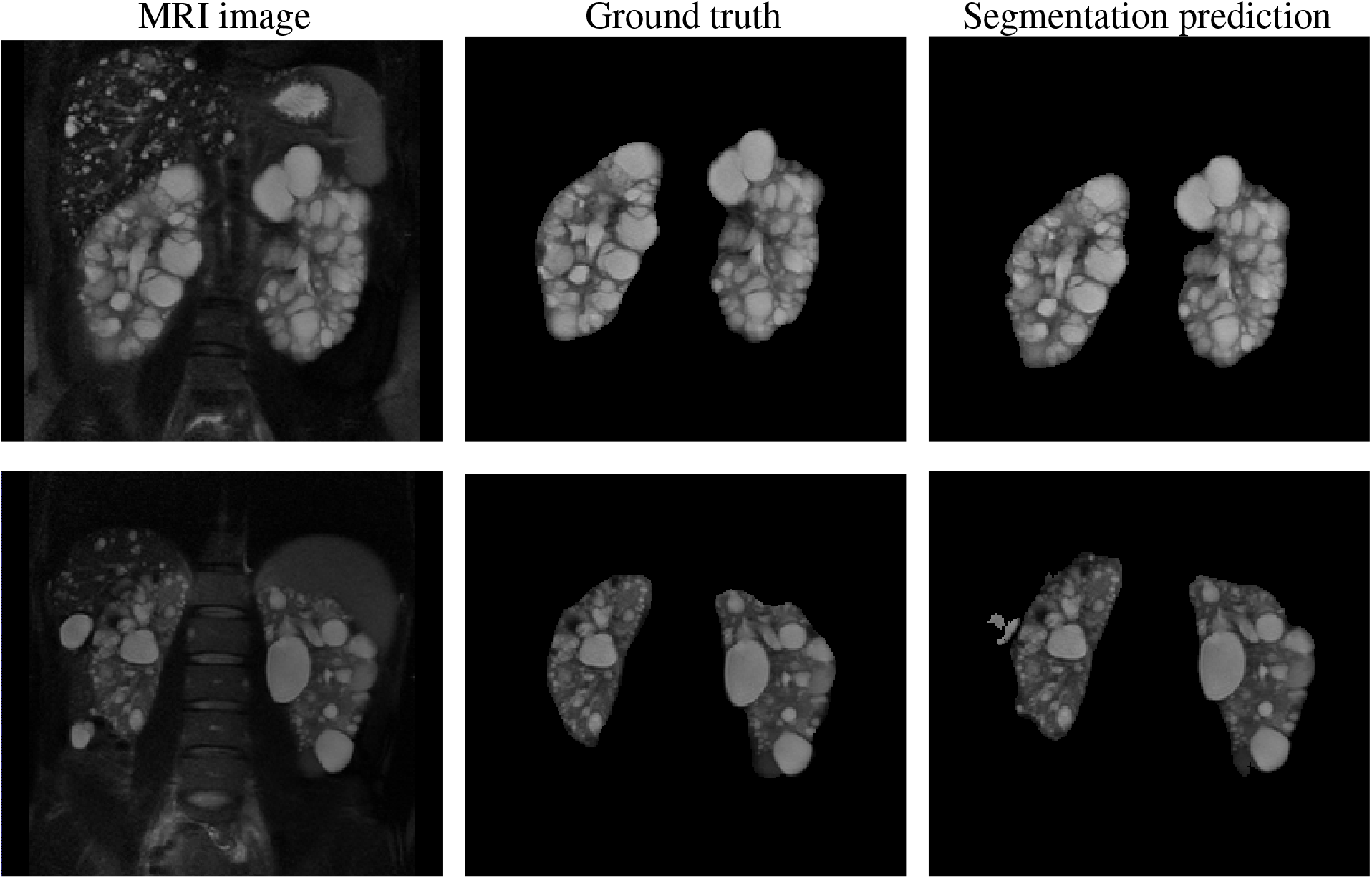
Kidney volume segmentation results. Left: T2-weighted MRI images, center: ground truth segmentation, and right: corresponding automatically segmented kidneys. Two examples show the segmented kidneys obtained from the segmentation network.

**Figure 5:**
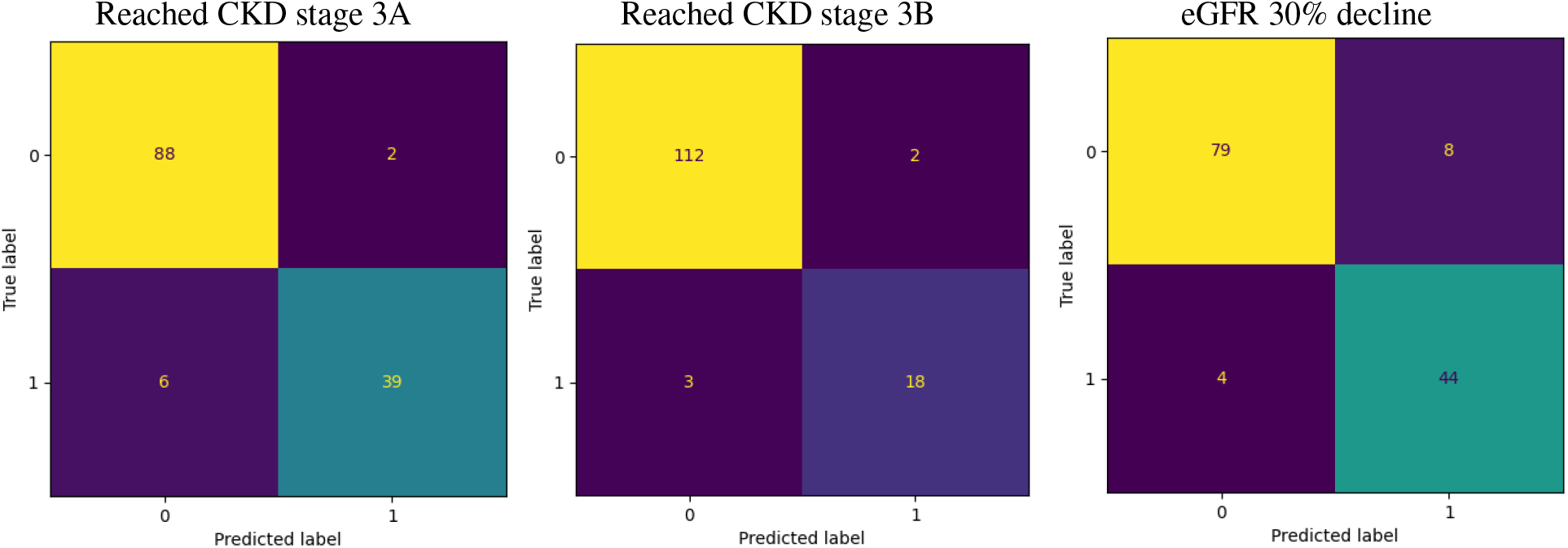
Confusion matrices depicting prognosis network predictions for reaching CKD stage 3A, reaching CKD stage 3B, and eGFR 30% decline from left to right, respectively.

**Figure 6:**
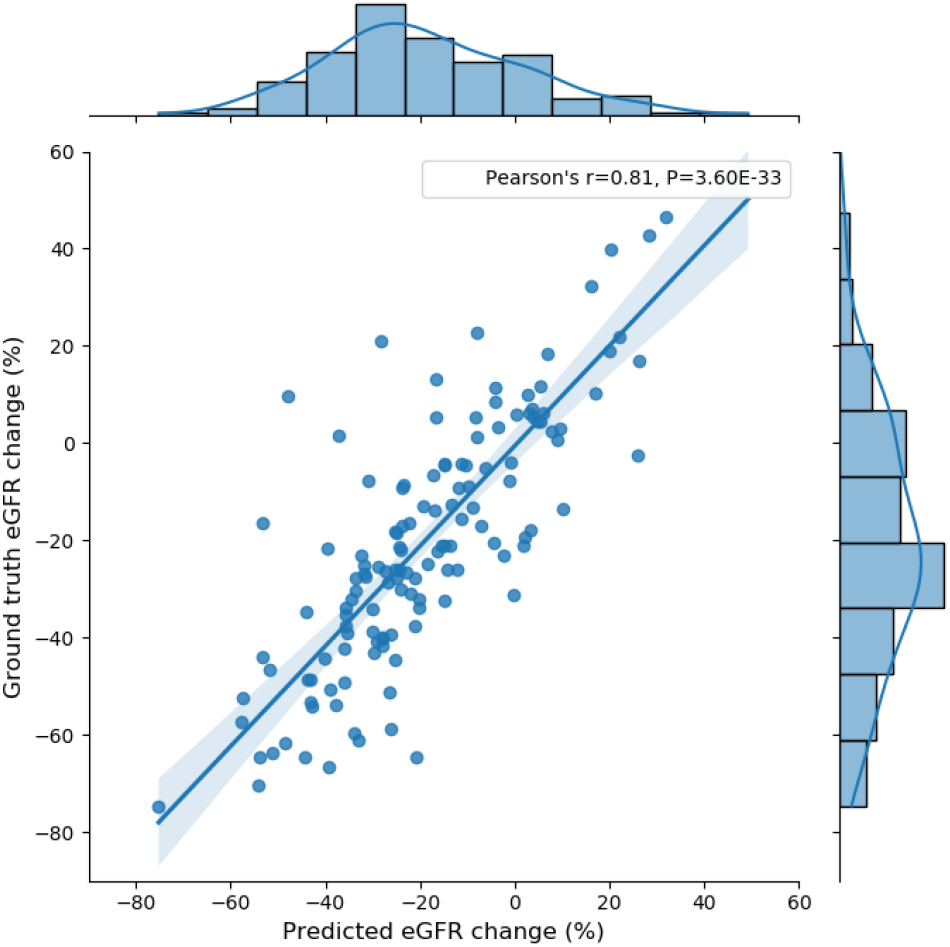
Correlation plot for predicted eGFR percent change v/s ground truth eGFR percent change after 8 years. The Pearson’s correlation coefficient is 0.81.

### 3.1. Kidney segmentation

The segmentation network segments kidney volumes with high accuracy as can be seen from the predicted HtTKV v/s ground truth HtTKV plot in figure 3. Here, Pearson’s correlation coefficient is 0.98 and the regression line has a coefficient of regression r^2^ value of 0.96. Furthermore, the mean percent difference between the predicted and the ground truth HtTKV is 13.47 ± 13.70 %. Figure 4 shows two examples of kidney segmentation from the segmentation network. Also, the Bland-Altman plot (see figure A.8) supports this finding depicting a bias of 66 ml/m. Only six samples exceed the 1.96 standard deviation range.

### 3.2. Classification/regression prognosis

The quantitative classification results are displayed in table 3, with the corresponding confusion matrices in figure 5. We observe that the number of positive samples is lower than the negative samples in each criterion (table 2 and figure 5).

The AUC for reaching CKD stage 3B is over 0.950 and on par with the corresponding value from the stateof-the-art (SOTA) method (0.960) [10]. However, the AUC of reaching CKD stage 3A is 0.965 and is higher than that of the SOTA method (0.940). Further, the 30% eGFR decline predictions reach an AUC of 0.952 and clearly exceed the results of the SOTA method (0.850). Furthermore, we observe that the precision and recall for each criterion is about 90%, indicating good performance of the classifiers (table 3).

Figure 6 depicts the predicted v/s ground truth eGFR percent change after 8 years. Here, Pearson’s correlation coefficient of 0.81 is attained (SOTA method = −0.700 [10]). The mean difference between predicted and ground truth eGFR percent change was found to be 1.12 ± 15.58 %. Plotting the respective Bland-Altman plot further supports this observation (see figure A.9). Most of the data is distributed within the 1.96 standard deviation range with a bias of only 1.12 percent change in eGFR, showing a small overestimation.

We further predicted eGFR values after 8 years for our patient data using the Mayo imaging classification tool [9, 21]. We found that the Pearson’s r for Mayo predicted eGFR was 0.64. In comparison, our prognosis network’s predicted values had an r value of 0.86 as shown in figure A.10 (Appendix A). Furthermore, the corresponding Bland-Altman plots show that the Mayo imaging classification tool underestimates the absolute predicted eGFR (bias of −1.76 ml/min/1.73 m^2^) while our method slightly overestimates the eGFR (bias of 1.18 ml/min/1.73 m^2^). Moreover, our approach has a smaller range of 1.96 standard deviations (−26.87 to 29.22 ml/min/1.73 m^2^) compared to the Mayo imaging classification tool (−44.88 to 41.37 ml/min/1.73 m^2^).

Finally, our model was also trained to predict each CKD stage distinctly for each patient after 8 years. Here, we reach a weighted f1-score of 0.851 (accuracy: 0.851) with an AUC of 0.972. The corresponding confusion matrix is shown in Fig. 7. As can be seen, most of the predictions are on the diagonal of the confusion matrix. Furthermore, the overall accuracy increases to 0.955 when we factor in misclassified samples from adjacent stages in the confusion matrix.

**Figure 7:**
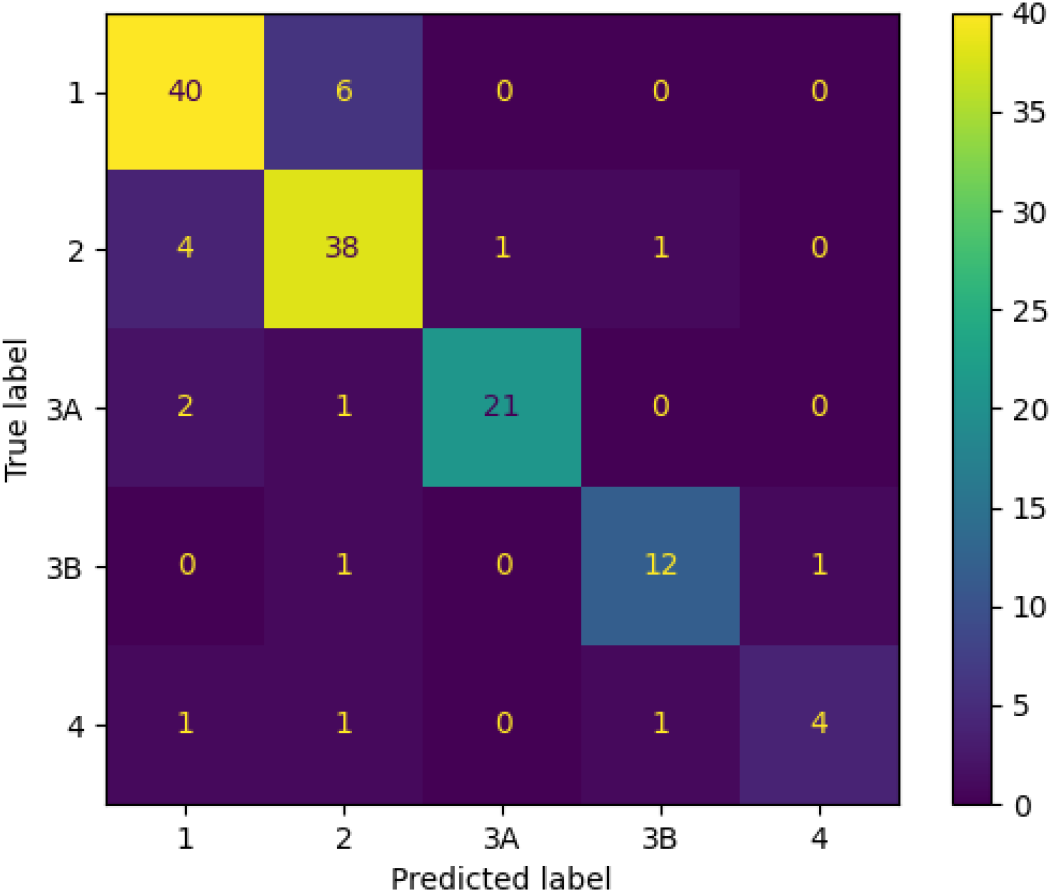
Confusion matrix depicting the predictions for distinct CKD stages after eight years. Weighted f1-score and AUC are 0.85 and 0.97, respectively.

## 4. Discussion

In this work, we combined T2-weighted MRIs of the kidneys with the established biomarkers (patient age, eGFR, and predicted HtTKV at the baseline visit) to predict renal function decline. Firstly, we segmented the kidneys automatically from the T2-weighted MR images using the attention U-Net and calculated the HtTKV from these segmentations. Afterwards, we used the segmented kidneys and the predicted HtTKV, age, and eGFR as inputs to our proposed prognosis network for prognosis. We found that our network performs at par with the state-of-the-art method [10] for classification of reaching CKD stage 3B. However, our approach achieves higher accuracy of prediction for reaching CKD stage 3A, 30% eGFR decline, and eGFR percent decline. We further extended our approach for classifying patients into CKD stages 1-4 after eight years.

### 4.1. Kidney segmentation

From a visual inspection, it can be seen that the kidney segmentation is accurate. However, when inspecting the predicted HtTKV vs ground truth HtTKV plot (figure 3), we observed that the network predicts slightly higher values, suggesting that it over-segments the kidneys. The mean percent difference of 13.47 ± 13.70 % between predicted and ground truth HtTKV confirms slight over-segmentation. A reason for this variability could be the slice thickness of 9.0 mm. Nonetheless, we still attained accurate prognosis performance with oversegmented kidneys as inputs to our model. Arguably, over-segmentation is better than under-segmentation since it is more likely to include the kidneys completely. Our segmentation network was originally established on T1 weighted MRIs and there we already showed good performance with respect to previously published studies [14]. In that study, we also noted that some samples contain cysts in various regions of the abdomen and less-defined kidney boundaries and shapes that might also lower the segmentation accuracy here. For comparison of T2 weighted segmentation, van Gastel et al. [22] achieved r^2^ value of 0.99 between the predicted and ground truth TKV segmentation. In comparison, our approach attains a lower r^2^ of 0.96, however, it is worth noting that van Gastel et al. [22] employed four times more data than our method.

### 4.2. Prognosis

The main objective of our proposed work was to automate ADPKD prognosis and obtain accurate results compared to the work of Kline et al. [10]. Often, we observe that tackling class imbalance is difficult for deep learning models[23, 24, 25], however, in this study, we achieved accurate results even though the number of positive samples was considerably less than negative samples, as seen in tables 1 and 2 (e.g. 23 positive samples v/s 112 negative samples for reaching CKD stage 3B). For the classification tasks (reaching CKD stage 3A, reaching CKD stage 3B, and 30% eGFR decline), our models obtain AUC > 0.950. Our method performs at par in the case of reaching CKD stage 3B classification when compared to the corresponding state-of-theart results from Kline et al. [10]. Here, we achieve an AUC of 0.957. In comparison, Kline et al. [10] achieve an AUC score of 0.960. It is worth noting that our dataset is slightly more imbalanced than the one used in [10] as depicted in table 2. Considering this imbalance, our approach is still robust enough in attaining accurate results for every criterion. Note-worthily, our method outperforms the state-of-the-art approach [10] in the cases of reaching CKD stage 3A, 30% eGFR decline, and eGFR percent change. For reaching CKD stage 3A and 30% eGFR decline, our AUCs of 0.965 and 0.952, respectively are higher than that of the stateof-the-art approach [10] (AUCs: 0.940 and 0.850, respectively).

Moreover, our model could be used to classify patients into the five CKD stages (1, 2, 3A, 3B, and 4) after eight years. Our network achieves an AUC and weighted f1-score of 0.972 and 0.851, respectively. We also find that about 95.5% of the predictions lie on the diagonal or the adjacent CKD stages in the confusion matrix (Fig. 7). However, the network misclassifies three cases in the CKD stage 4 category. This occurs due to the class imbalance in the CKD stage 4 category as it consists of only seven cases. Increasing the dataset by including more samples for this class might improve the model’s performance.

The regression task of eGFR percent change reaches a Pearson’s r value of 0.810 as compared to −0.700 of the state-of-the-art method [10]. The features in [10] are negatively correlated to the eGFR percent change, hence, the negative sign. However, our method has a positive correlation since we directly compare the predicted eGFR percent change with the ground truth eGFR percent change (figure 6). Furthermore, the residual standard deviation of our approach was also smaller (15.584 % vs 17.900 %). It is clear that the networks are more accurate in predicting eGFR change (classification or regression) after 8 years. Compared to the Mayo imaging classification tool [9], our algorithm reached a higher correlation of the predicted eGFR to the ground truth eGFR (r=0.64 vs. r=0.86, figure A.10).

The main advantages of our approach over the one presented by Kline et al. [10] are automation and time consumption. Kline et al. [10] segmented the kidneys manually, which is time-consuming and operator-dependent. Meanwhile, our approach segments whole kidney volumes automatically in a few seconds. Furthermore, Kline et al. [10] used three feature values (energy, entropy, and correlation) from one MRI, in contrast, we captured more features from each kidney volume that are helpful in robust prognosis. Furthermore, our model could also classify patients into different CKD stages. This classification might better support the diagnosis of patients with ADPKD since it allows for precisely predicting the change in CKD class and therefore, the decline in renal function over time rather than just predicting if a person will reach a certain CKD stage or not.

A few other works exist that predict renal function decline but are not directly comparable to our approach because of differences in datasets and renal function decline definitions. Cao et al. [26] compared various machine learning algorithms on a dataset of 2166 subjects. They defined renal function decline as eGFR decline of more than 3 ml/min/1.73 m^2^/year or follow-up eGFR < 60 ml/min/1.73 m^2^ (i.e., CKD stage 3A to 5). The algorithms were trained using 24 predictive variables, e.g., age and gender. Their best-performing algorithm was gradient boosting, reaching an AUC of 0.914 on the test data. Zhao et al. [27] explored a prediction model to predict CKD stage 3A or positive proteinuria in a cohort of 348 subjects. Their prediction model combined a genetic risk score model with a non-genetic risk score model by incorporating genetic and non-genetic factors for prediction. This combined model achieved an AUC value of 0.894.

There exist a few approaches that seek to classify distinct CKD stages automatically. However, all approaches do not incorporate imaging data nor do a longterm (larger than 18 months,[28]) prediction of renal function decline. Debal and Sitote [29] created a random forest model that predicts 5 CKD stages (from 1 to 5). They achieved an f1-score of 0.778 with a dataset of 1718 samples. Ilyas et al. [30] employed a dataset of 400 instances and predicted 6 CKD stages (stages 3A and 3B separately). They obtained an overall accuracy of 0.855 using the J48 algorithm, a decision tree based approach. Finally, Rady and Anwar [31] trained a network of probabilistic neural networks on a dataset of 361 patients, achieving an overall accuracy of 0.967. Despite the difference in the underlying data like medical records (such as age, blood pressure, hypertension, hemoglobin, etc.) our integrated approach incorporating imaging information and clinical information (age, eGFR) could achieve a similar or higher classification accuracy while employing only about 3-fold fewer data and allowing for accurate kidney segmentation and TKV estimation simultaneously.

The main limitation of our approach pertains to the transfer of the deep learning models to other related datasets. Since MRI sequences can vary among various sites and vendors, it usually makes the models less generalizable. Our models will generalize well if the MRI protocol is similar to the one used by the NIDDK-CRISP study. Fine-tuning the networks on new datasets might be helpful, however, that will require at least tens of new subjects. Furthermore, several approaches exist that help adapt networks to new datasets [32, 33, 34, 35, 36, 37], however, this is a subject of future work since our main focus was to show the feasibility of an automated prediction. Another shortcoming is that the proposed method is only applicable to a time interval of 8 years since we could only compare it directly with the state-of-the-art method. Prediction of kidney function decline over other time intervals will be a future task.

In the future, we plan to use T1-weighted kidney volumes and combine them with their T2-weighted counterparts to extract better features and attempt to improve performance. Furthermore, we plan to do multi-task learning by combining the training of segmentation and prognosis networks.

In conclusion, we have presented an automated approach to predict disease progression in ADPKD, in terms of eGFR decline and CKD stage change by integrating imaging information and clinical data. Simultaneously, renal segmentations are also obtained and used in further diagnostic tasks. Our approach might improve monitoring and support the prognosis of ADPKD patients from the earliest disease stages.

## Data Availability

All data produced are available upon request online at https://repository.niddk.nih.gov/home/

## 6. Acknowledgments

The Consortium for Radiologic Imaging Studies of Polycystic Kidney Disease (CRISP) was conducted by the CRISP Investigators and supported by the National Institute of Diabetes and Digestive and Kidney Diseases (NIDDK). The data from the CRISP study reported here were supplied by the NIDDK Central Repository. This manuscript was not prepared in collaboration with investigators of the CRISP study and does not necessarily reflect the opinions or views of the CRISP study, the NIDDK Central Repository, or the NIDDK. We are thankful to the NIDDK for providing us with the patient data from the CRISP study. We gratefully acknowledge the support of the NVIDIA Corporation with the donation of an NVIDIA Titan Xp used for this research. This research project is part of the Research Campus M2OLIE and funded by the German Federal Ministry of Education and Research (BMBF) within the Framework “Forschungscampus: public-private partnership for Innovations” under the funding code 13GW0388A. This study was supported in part by the German Federal Ministry of Education and Research (BMBF) under the funding code 01KU2102, and the Italian Ministry of Health, under the frame of ERA PerMed (ERAPERMED2020-326 - RESPECT). Dr. Caroli acknowledges a grant from the Italian Association for Polycystic Kidney (Associazione Italiana Rene Policistico - AIRP). The authors wish to thank Dr Norberto Perico, Istituto di Ricerche Farmacologiche Mario Negri IRCCS, Bergamo, Italy, for his valuable comments and suggestions.

## Appendix A. Comparison of the Mayo Image classification tool to the prognosis network

Figures A.8 and A.9 depict the Bland-Altman plots comparing our models’ prediction for HtTKV and eGFR percent change against the corresponding ground truth values.

**Figure A.8:**
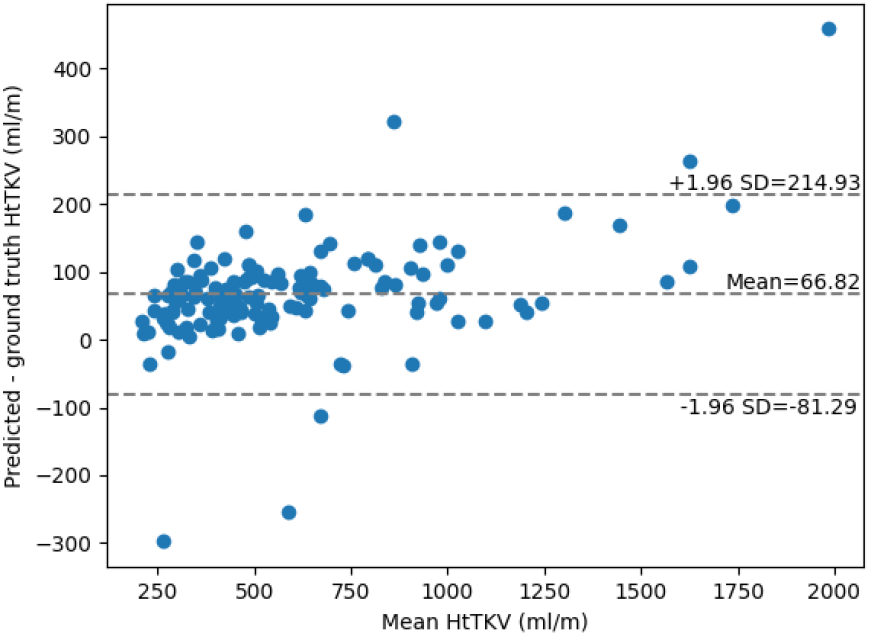
Bland-Altman plot comparing model predicted HtTKV (ml/m) values to the ground truth values.

**Figure A.9:**
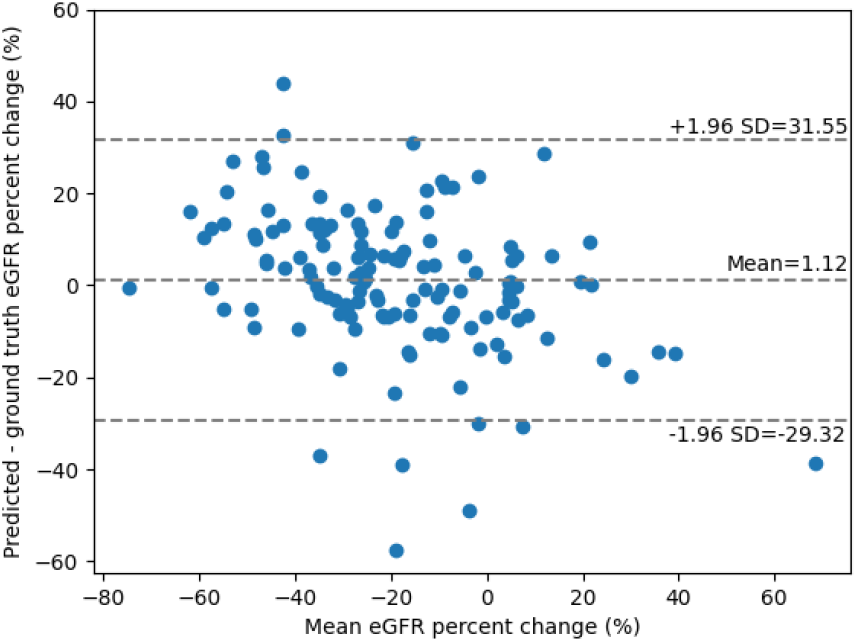
Bland-Altman plot comparing model predicted eGFR percent change (%) values to the ground truth values.

Figure A.10 shows the correlation plots for eGFR predictions by the Mayo Image classification tool and by the prognosis network. The Mayo Image classification tool eGFR predictions have a Pearson’s r of 0.64, in comparison, our network attains an r value of 0.86. Furthermore, it shows the Bland-Altman plots for comparison. The standard deviation for our model is lower than the Mayo Image classification tool.

**Figure A.10:**
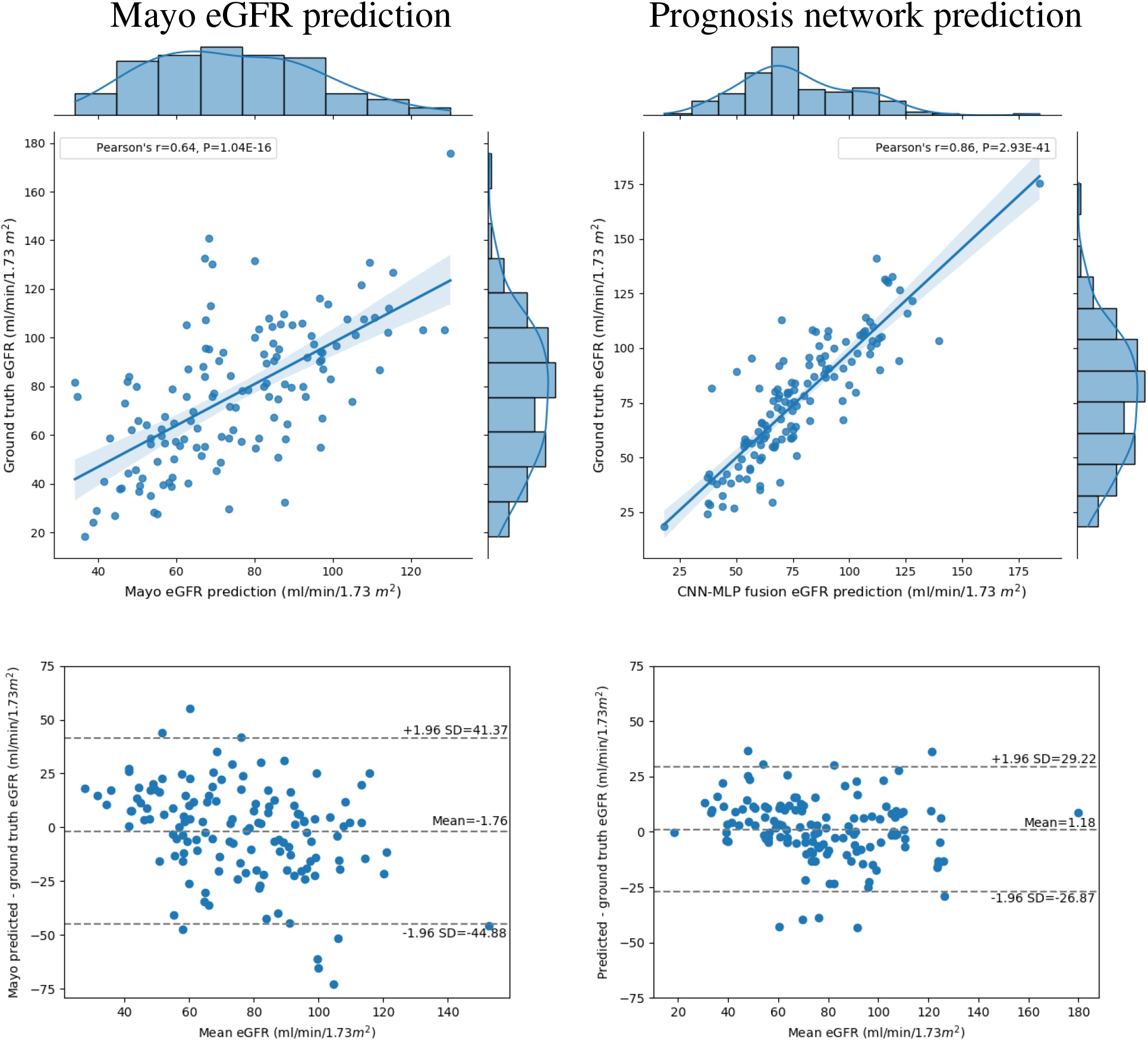
Correlation plots showing predicted v/s ground truth eGFR values after 8 years in the first row. The left plot is obtained from Mayo classification tool [9, 21] and the right plot is from our described prognosis network. The second row depicts the corresponding Bland-Altman plots.

